# HIV Incidence Could Rise by 68% in 11 States if Ryan White Ends: A Simulation Study

**DOI:** 10.1101/2025.07.31.25332525

**Authors:** Melissa Schnure, Ryan Forster, Joyce L. Jones, Catherine R. Lesko, D. Scott Batey, Isolde Butler, Dafina Ward, Karen Musgrove, Keri N. Althoff, Mamta K. Jain, Kelly A. Gebo, David W. Dowdy, Maunank Shah, Parastu Kasaie, Anthony T. Fojo

## Abstract

**Objectives:** To estimate the increase in HIV infections in 11 US states if Ryan White services are interrupted or ended.

**Methods:** We applied a population-level model of HIV transmission to 11 states. We represented the proportion of people with HIV receiving Ryan White AIDS Drug Assistance, Outpatient Health services, or Support services, and simulated a loss of suppression in each category if services permanently end or return after delays of 1.5 or 3.5 years.

**Results:** Cessation of Ryan White services in 2025 was projected to result in 69,695 additional infections from 2025-2030 (95% credible interval 18,943 to 123,628) – 68% (18% to 118%) more than if Ryan White were continued. Temporary interruptions of 1.5 and 3.5 years resulted in 26,951 (7,341 to 47,534) and 53,594 (14,645 to 94,860) additional infections, respectively. Excess infections varied across states, from a 45% increase in Texas to 126% in Missouri.

**Conclusions:** Projected increases in HIV infections due to disruptions of Ryan White services threaten the progress made in curtailing the US HIV epidemic, illustrating the critical role Ryan White plays in preventing HIV transmission.

## Introduction

Enacted in 1990, the Ryan White HIV/AIDS Program provides comprehensive services for over half of the 1.2 million people with HIV (PWH) in the US and often functions as a “payer of last resort” for those who would otherwise be unable to access care^1^. The program provides assistance through several medical and support mechanisms: payment for antiretroviral therapy through the AIDS Drug Assistance Program, direct organizational funding for routine outpatient ambulatory health services, and non-medical support including case management or transportation^2^. As a result, Ryan White clients have high rates of viral suppression, rendering those clients unable to transmit HIV^2–4^.

The Ryan White program is administered by the Health Resources and Services Administration^1^. It operates under annual budgetary approval from Congress and is distributed through grants to local jurisdictions and HIV care organizations^1^. Disruptions to Ryan White services could substantially increase HIV transmission in the US. Using a mathematical HIV transmission model, we sought to estimate the increase in HIV infections in 11 states if Ryan White services were interrupted or discontinued.

## Methods

The Johns Hopkins Epidemiologic and Economic Model is a dynamic, compartmental model of HIV transmission that stratifies the adult population by age, race/ethnicity, sex, and HIV status^5^. Model calibration has been described previously^5^; we simulated HIV epidemics at the state level using a similar process (see Supplement).

We simulated HIV epidemics from January 2025 to December 2030 in 11 states, in which 63% of people diagnosed with HIV in the US reside: Alabama, California, Florida, Georgia, Illinois, Louisiana, Mississippi, Missouri, New York, Texas, and Wisconsin. These states were chosen to represent varied geographic regions, Medicaid expansion status, and prioritization within the national *Ending the HIV Epidemic* initiative^6^.

A prior analysis examining the impact of Ryan White program cessation at the city level describes our methodology for representing program services^7^. In short, we simulated the proportion of PWH receiving services, dividing them into three mutually exclusive categories: (1) AIDS Drug Assistance (anyone receiving drug assistance, regardless of whether they also received other services), (2) Outpatient Ambulatory Health Services at HIV care facilities (excluding those receiving drug assistance), and (3) other Ryan White support services such as case management or transportation (but not drug assistance or Outpatient Ambulatory Health Services)^2^. Calibration targets for Ryan White service utilization included the number of clients accessing each service type and the proportion of each client type who were virally suppressed in each state (see Supplement).

Based on a survey of Ryan White clinic directors, administrators, and public health officials who estimated the proportion of clients who would lose viral suppression if services stopped, we assumed a mean 65% (interquartile range 40-90%) loss of viral suppression among recipients of AIDS Drug Assistance, 49% (25-70%) among other outpatient health services recipients, and 37% (10-60%) among Ryan White clients receiving other services^7^. We ran 1,000 simulations in each state, sampling effects of ending Ryan White services from the range of survey responses, conditional on state Medicaid expansion status. As survey respondents might overestimate the impact of disruptions, we performed a “conservative” secondary analysis in which we also incorporated data from two published studies on the association between Ryan White services and viral suppression^7–9^.

We modeled three scenarios: (1) “Cessation,” in which services permanently stop in July 2025 and viral suppression drops without recovery; and two scenarios in which services stop in July 2025 but viral suppression begins to recover in either (2) January 2027 (“Brief [1.5-year] Interruption”) or (3) January 2029 (“Prolonged [3.5-year] Interruption”).

## Results

Assuming Ryan White services continue, we projected 103,101 new HIV infections (95% credible interval 97,773 to 109,048) from 2025-2030 across all 11 states. If Ryan White services end permanently in 2025 (“Cessation”), we projected 69,695 additional infections (18,943 to 123,628) from 2025-2030, an excess of 68% (18 to 118%, Figure 1 and Supplemental Figures S3-4). Projected numbers of additional infections were lower in other scenarios. In the “Brief [1.5-year] Interruption” scenario, we projected 26,951 (7,341 to 47,534) additional infections, an excess of 26% (7 to 46%). In the “Prolonged [3.5-year] Interruption” scenario, we projected 53,594 (14,645 to 94,860) additional infections, an excess of 52% (14 to 90%). In the “Conservative” analysis, the number of additional infections were approximately half of those from the primary analysis for all scenarios (Supplemental Figures S5-6).

**Figure 1:**
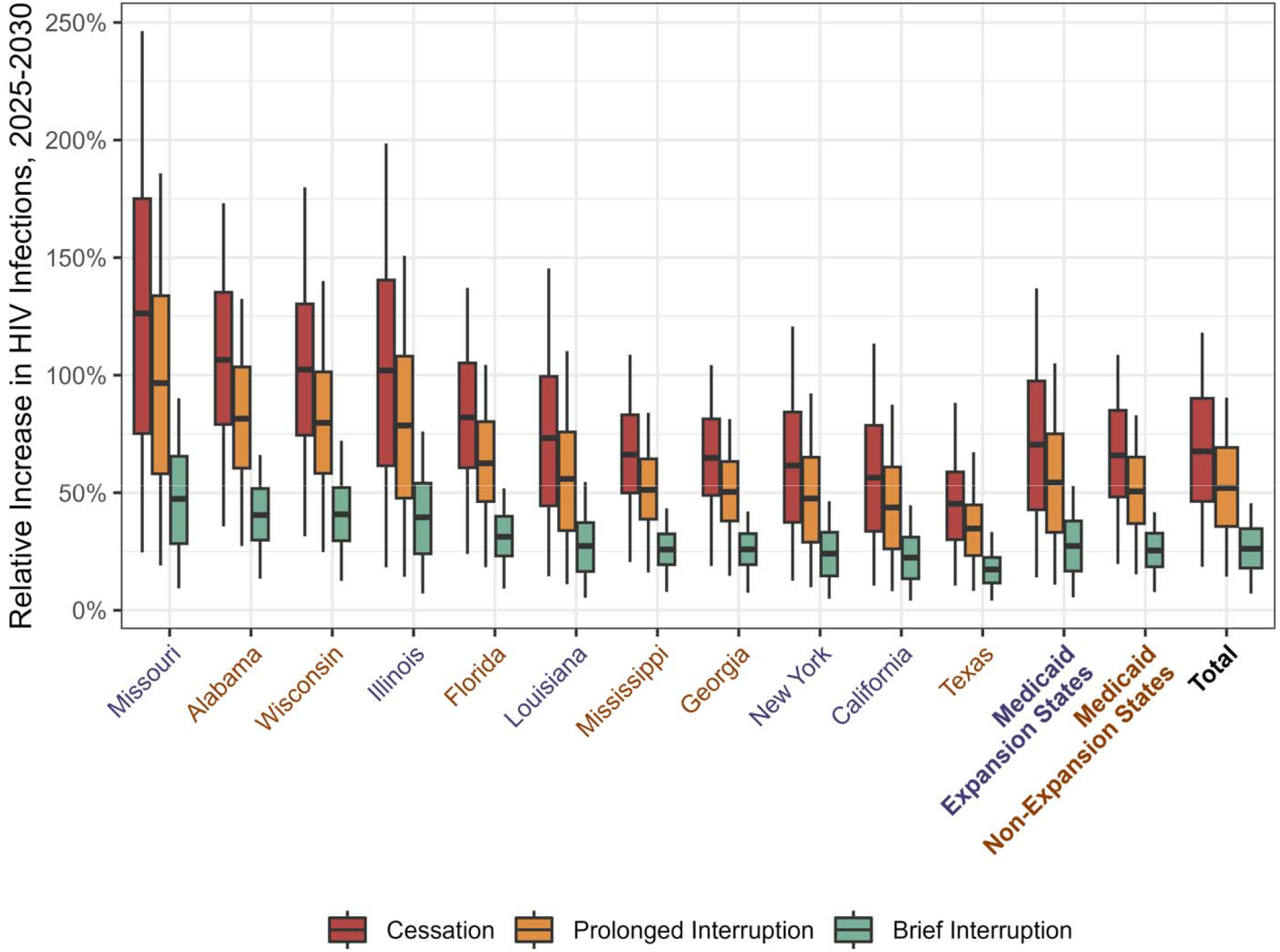
State-Level Relative Excess HIV Infections from 2025-2030 If Ryan White Programs are Stopped or Interrupted. Boxplots display the projected percentage increase in new HIV infections under three scenarios in which all Rya White services stop in July 2025: “Cessation” (red) – viral suppression among Ryan White clients never recovers; “Prolonged [3.5-year] Interruption” (orange) – viral suppression among Ryan White clients recovers from January t December 2029; “Brief [1.5-year] Interruption” (green) – viral suppression among Ryan White clients recovers from January to December 2027. The value along the y-axis represents the relative increase in cases vs. a scenario where Ryan White services continue uninterrupted. The dark horizontal lines indicate the median projection across 1,000 simulations, the boxes indicate interquartile ranges (IQR), and whiskers cover the 95% credible interval. Medicai non-expansion states are denoted by orange labels; expansion states are indicated by purple ones.

The expected increases in HIV infections due to program cessation varied across states (Figure 1), from 45% (10 to 88%) in Texas to 126% (25 to 246%) in Missouri. In four states—Missouri, Alabama, Wisconsin, and Illinois—the projected number of infections would more than double due to program cessation (i.e., an excess greater than 100% compared to program continuation). See https://jheem.org/ryan-white-state-level for state-level results. In a secondary analysis exploring between-state variation, we found that states with a greater proportion of PWH receiving Ryan White services in 2025 had higher levels of excess HIV infections (Supplemental Figure S7). Across all states combined, infections increased most among adults under 35 years old and men who have sex with men, without large differences by race (Supplemental Text S1).

## Discussion

We used a validated HIV transmission model, informed by CDC surveillance data and Ryan White Program reports, to project the impact of disruptions to the US Ryan White HIV/AIDS Program in 11 US states from 2025-2030. Permanently ending these services would lead to nearly 70,000 additional HIV infections in the next five years. However, even temporary interruptions could result in a considerable rise of over 26,000 additional infections. Impacts are expected to vary substantially at the state level, with the largest increases in new HIV infections in Missouri, Alabama, Wisconsin, and Illinois.

Our analysis is subject to limitations, especially in that the individual-level effects of ending Ryan White services are uncertain. Without empirical evidence on these effects, we relied on estimates from those with expertise in Ryan White programming. We attempted to capture both the uncertainty around these estimates by simulating effects across the range of survey responses, and any potential bias in overestimating effects by performing a secondary, “conservative” analysis. Second, we assume that cessation or interruption of programming will only affect individuals accessing services. It is likely, however, that non-Ryan White clients would also experience impacts of funding cuts to clinics. From this perspective, our estimates may underestimate the true impacts of reducing Ryan White funding. Finally, while we limited the scope of our analysis to estimate impacts on future transmission, interrupting Ryan White services would also impact both quality of life and mortality for those currently living with HIV.

The modeling approach used here has several strengths. Our Bayesian calibration process is a rigorous approach to handling unknown variables in HIV epidemiology. Furthermore, by calibrating to stratified epidemiologic data at the state level, we can compare across settings and capture local differences in HIV epidemics.

In summary, projected increases in new HIV infections through the loss of viral suppression threaten progress made in curtailing the US HIV epidemic, should Ryan White services be interrupted or discontinued. These findings illustrate the critical role that Ryan White services play in preventing the transmission of HIV and the importance of maintaining these services at the state level.

## Data Availability

All data produced in the present study are available upon reasonable request to the authors

https://jheem.org/ryan-white-state-level

