## Supplement for "HIV Incidence Could Rise by 68% in 11 States if Ryan White Ends: A Simulation Study"

### **Technical Supplement**

### **Model Structure**

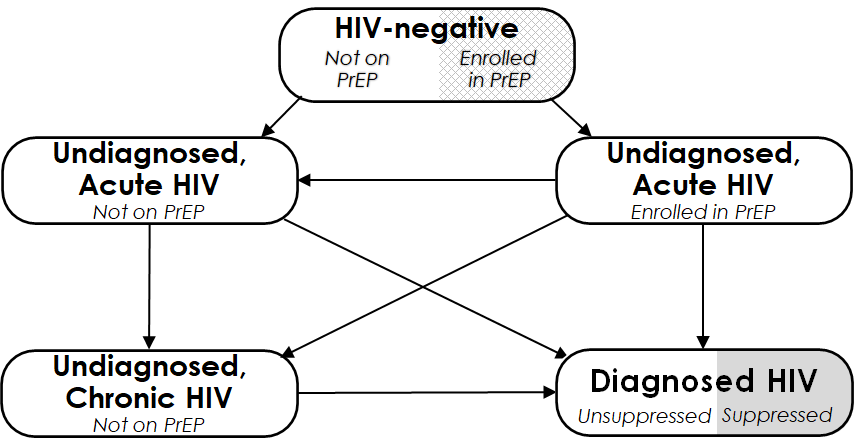

**Figure S1: Model structure.** This figure depicts the compartments representing HIV status. Each of the five compartments is further stratified by age (13–24, 25–34, 35–44, 45–54, and ≥55 years), race/ethnicity (Black, Hispanic, and other), sex and sexual behavior (female, heterosexual male, and men who have sex with men (MSM), and intravenous drug use history (never used, active use, and prior use). “Acute HIV” refers to the first 2.9 months following infection, during which risk of transmission is high.

### **Ryan White Services**

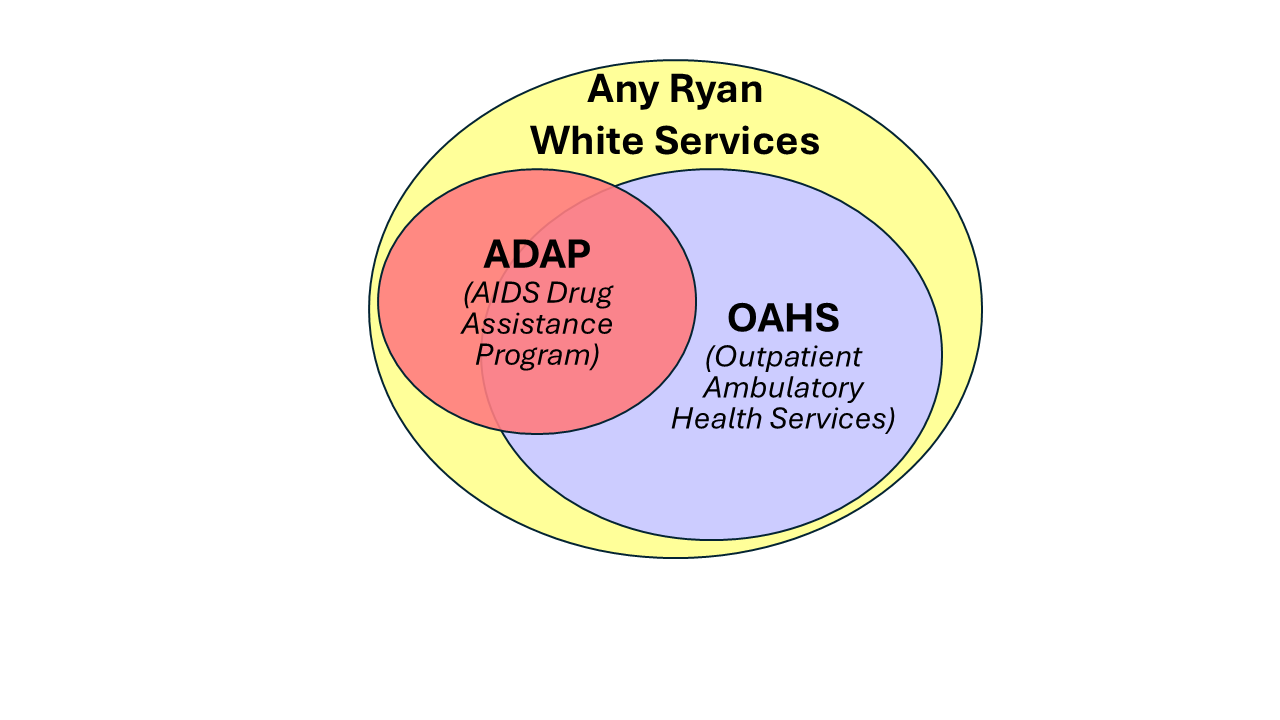

**Figure S2: Venn-diagram of Ryan White Services.** This depicts the mutually exclusive funding groups explored in these analyses. (1) AIDS Drug Assistance Program grants may be used to pay for antiretroviral therapy directly, pay premiums for health insurance, and help with medication copays; (2) Outpatient Ambulatory Health Services provides direct funding to HIV care facilities for outpatient clinical care; and (3) Other Ryan White services consist primarily of non-medical support services, such as case management, transportation assistance, adherence support, and housing assistance.

### **Calibration Targets**

| **Outcome Target** | **Years** | **Stratification** | **Source** |
| --- | --- | --- | --- |
| Proportion receiving AIDS Drug Assistance (ADAP) | 2017–2021 | Age, Sex, Race,  Risk Factor | Ryan White AIDS Drug Assistance Program Annual Client-Level Data Reports (HRSA) |
| Viral Suppression among ADAP Clients | 2017–2023 | Total | National Ryan White HIV/AIDS Program Part B & ADAP Monitoring Project Annual Report (NASTAD) |
| Non-ADAP RWHAP Clients | 2017–2023 | Age, Sex, Race,  Risk Factor | Ryan White HIV/AIDS Program Services Annual Reports |
| Outpatient Ambulatory Health Services (OAHS) Clients | 2017–2023 | Age, Race | Ryan White HIV/AIDS Program Services Annual Reports |
| Viral Suppression among OAHS Clients | 2017–2023 | Age, Race | Ryan White HIV/AIDS Program Services Annual Reports |

**Table S1: Calibration targets for Ryan White-specific parameters.**

| **Outcome Target** | **Years** | **Stratification** | **Source** |
| --- | --- | --- | --- |
| Diagnosed HIV Prevalence | 2008–2023 | Age, Sex, Race,  Risk Factor | CDC Surveillance Reports, AtlasPlus |
| New HIV Diagnoses | 2008–2023 | Age, Sex, Race,  Risk Factor | CDC Surveillance Reports, AtlasPlus |
| HIV Suppression | 2008–2023 | Age, Sex, Race,  Risk Factor | AtlasPlus |
| HIV Mortality (all-cause among PWH) | 2008–2023 | Sex | CDC Surveillance Reports, AtlasPlus |
| General Mortality | 2007–2019 | Total | US Census |
| AIDS Diagnoses | 1985–1993 | Sex, Race Risk Factor | CDC Surveillance Reports, CDC WONDER |
| Awareness of HIV Status | 2008–2023 | Total | AtlasPlus |
| Proportion Tested for HIV | 2010–2023 | Age, Sex, Race,  Risk Factor | BRFSS |
| Positivity among CDC-funded HIV Tests | 2014–2020 | Total | CDC Surveillance Reports |
| Number of People Prescribed PrEP | 2007–2023 | Age, Sex, Race | AIDSVu, AtlasPlus |
| PrEP Indications | 2017–2018 | Age, Sex | AtlasPlus |
| Proportion Using Heroin | 2008–2023 | Age | NSDUH |
| Proportion Using Cocaine | 2008–2023 | Age | NSDUH |
| Immigration | 2011–2023 | Age, Race, Sex | American Communities Survey |
| Emigration | 2011–2023 | Age, Race, Sex | American Communities Survey |
| Population | 2010–2023 | age, sex, race | US Census |

**Table S2: Calibration targets for Base JHEEM model.**

### **Additional Results and Secondary Analyses**

**Supplemental Text S1: Results by subgroup**

By subgroup, the main differences in the relative increase in infections were observed across strata of age and risk group. The youngest age group (age 13-24) saw a 79% increase (95% credible interval 22 to 138%) in infections from 2025-2030, compared to 64% (17 to 112%) across all other ages. By risk group, there was an 85% (23 to 147%) increase among men who have sex with men (MSM) compared to a 48% (13 to 85%) increase for non-MSM. There were no notable differences in the relative increase by race: 68% (109 to 117%) among Black residents, 66% (17 to 118%) among Hispanic residents, and 68% (18 to 122%) among residents of other races.

**Figure S3: State-Level Excess HIV Infections from 2025-2030 If Ryan White Programs are Stopped or Interrupted**

|  | **Continuation** | **Cessation** | | **Prolonged Interruption** | | **Brief Interruption** | |
| --- | --- | --- | --- | --- | --- | --- | --- |
| **State** | Number of  Incident Infections | Number of Excess Infections | Relative Excess Infections* | Number of Excess Infections | Relative Excess Infections* | Number of Excess Infections | Relative Excess Infections* |
| Missouri | 2,826 | 3,557 | 126% | 2,722 | 97% | 1,334 | 47% |
|  | [2,413-3,222] | [707-6,991] | [25-246%] | [558-5,259] | [19-186%] | [271-2,557] | [9-90%] |
| Alabama | 3,818 | 4,063 | 106% | 3,109 | 81% | 1,544 | 40% |
|  | [3,327-4,261] | [1,330-6,788] | [36-173%] | [1,030-5,183] | [27-132%] | [503-2,598] | [13-66%] |
| Wisconsin | 1,442 | 1,461 | 102% | 1,137 | 80% | 582 | 41% |
|  | [1,145-1,755] | [464-2,471] | [31-180%] | [361-1,926] | [25-140%] | [183-991] | [12-72%] |
| Illinois | 5,105 | 5,201 | 102% | 4,009 | 79% | 2,016 | 40% |
|  | [4,356-6,203] | [958-10,252] | [18-198%] | [744-7,833] | [14-151%] | [370-3,930] | [7-76%] |
| Florida | 19,222 | 15,767 | 82% | 12,016 | 63% | 6,006 | 31% |
|  | [16,909-22,100] | [4,576-26,938] | [24-137%] | [3,531-20,344] | [18-104%] | [1,777-10,103] | [9-52%] |
| Louisiana | 4,545 | 3,315 | 73% | 2,531 | 56% | 1,238 | 27% |
|  | [3,825-5,288] | [643-6,712] | [14-145%] | [492-5,121] | [11-110%] | [236-2,547] | [5-55%] |
| Mississippi | 2,565 | 1,689 | 66% | 1,306 | 51% | 658 | 26% |
|  | [2,177-3,017] | [514-2,690] | [21-109%] | [399-2,085] | [16-84%] | [193-1,070] | [8-43%] |
| Georgia | 12,198 | 7,886 | 65% | 6,127 | 50% | 3,150 | 26% |
|  | [10,687-14,419] | [2,300-12,775] | [19-104%] | [1,796-9,899] | [15-81%] | [914-5,092] | [7-42%] |
| New York | 10,563 | 6,499 | 62% | 5,022 | 48% | 2,542 | 24% |
|  | [9,025-12,200] | [1,254-13,125] | [13-121%] | [980-9,962] | [10-92%] | [497-5,032] | [5-46%] |
| California | 16,519 | 9,275 | 56% | 7,188 | 44% | 3,679 | 22% |
|  | [14,276-19,355] | [1,758-18,334] | [10-113%] | [1,369-14,176] | [8-87%] | [681-7,228] | [4-45%] |
| Texas | 24,299 | 10,982 | 45% | 8,428 | 35% | 4,201 | 17% |
|  | [21,391-28,267] | [2,614-21,893] | [10-88%] | [2,010-16,680] | [8-67%] | [994-8,228] | [4-33%] |
| **Medicaid Expansion States** | 39,558 | 27,847 | 70% | 21,471 | 54% | 10,809 | 27% |
|  | [36,801-42,823] | [5,487-53,565] | [14-137%] | [4,277-41,100] | [11-105%] | [2,165-20,720] | [5-53%] |
| **Medicaid Non-Expansion States** | 63,543 | 41,848 | 66% | 32,123 | 51% | 16,142 | 25% |
|  | [59,156-68,774] | [12,862-69,674] | [20-109%] | [9,874-53,208] | [15-83%] | [4,925-26,618] | [8-42%] |
| **Total** | 103,101 | 69,695 | 68% | 53,594 | 52% | 26,951 | 26% |
|  | [97,773-109,048] | [18,943-123,628] | [18-118%] | [14,645-94,860] | [14-90%] | [7,341-47,534] | [7-46%] |
|  |  | 0% \| 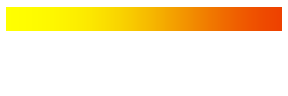 \| 100% | | | | | |

The “Continuation” column gives the mean and 95% credible interval, across 1,000 simulations, projected incident HIV infections from 2025-2030 if Ryan White programs continue uninterrupted. The columns labeled “Number of Excess Infections” give the mean and 95% interval of the absolute number of excess HIV infections expected from 2025-2030 under three scenarios where Ryan White programs are stopped in July 2025: “Cessation” (viral suppression among Ryan White clients never recovers), “Prolonged Interruption” (viral suppression recovers from January to December 2029), and “Brief Interruption” (viral suppression recovers from January to December 2027). The columns labeled “Relative Excess Infections” give the percent change in projected incident infections, relative to “Continuation”. Cells are shaded according to the relative excess infections. Medicaid non-expansion states are denoted by orange text; expansion states are denoted by purple text.

**Figure S4: Projected HIV Incidence if Ryan White Programs End or are Interrupted**

**Cessation**

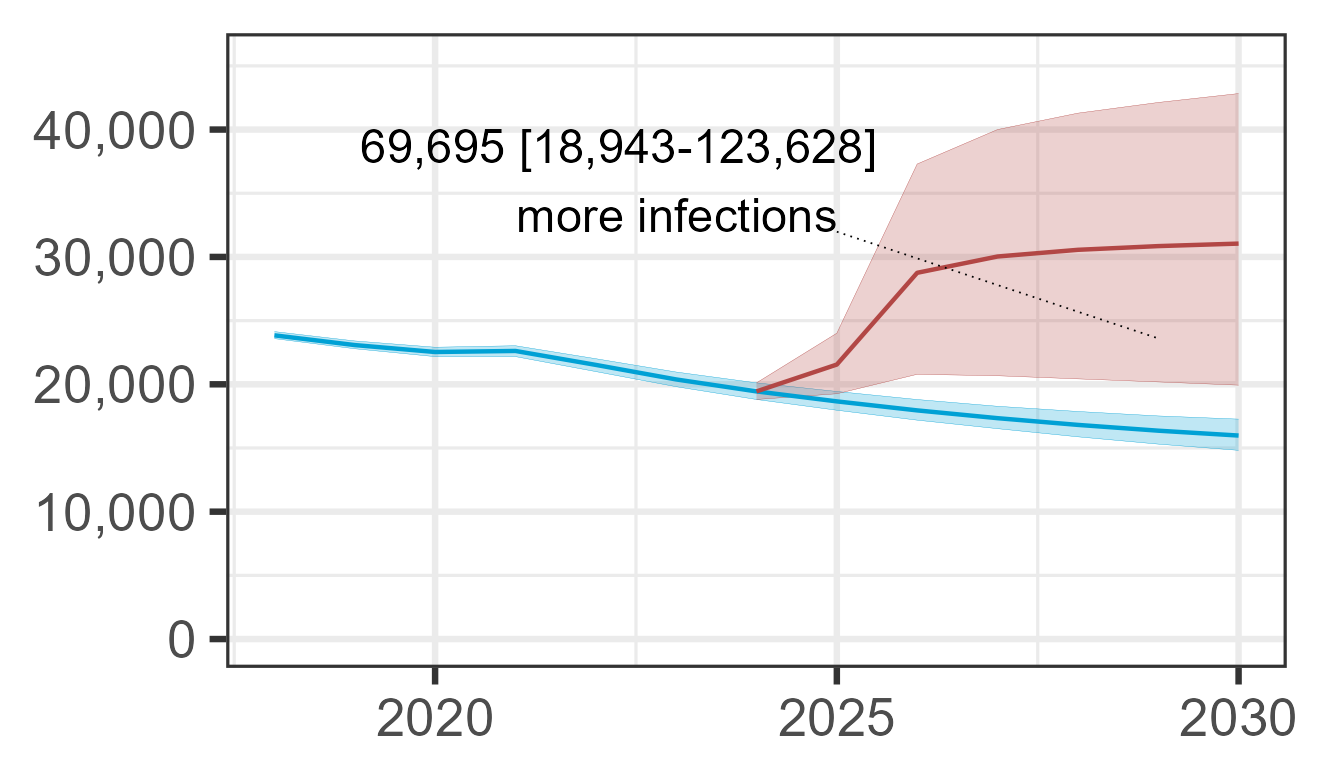

**Prolonged Interruption**

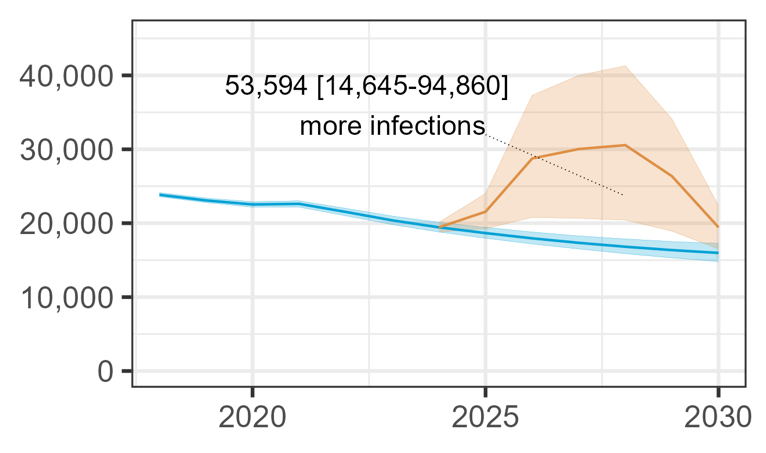

**Brief Interruption**

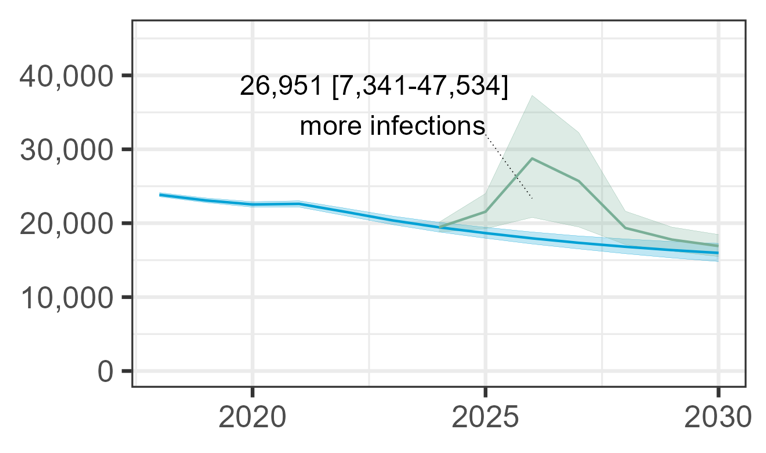

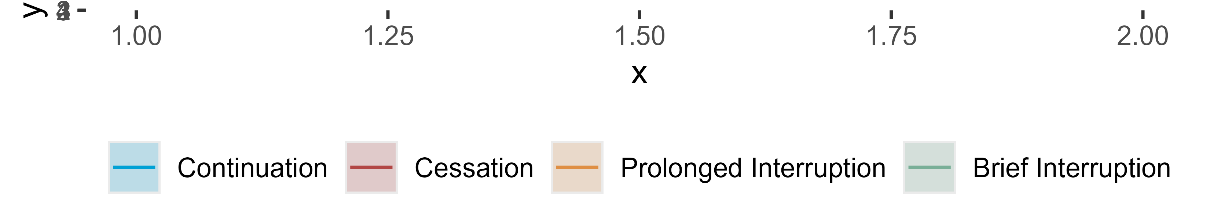

Projection across all 11 states. Y-axes give the projected number of infections. Lines denote the mean across 1,000 simulations; ribbons give the 95% credible interval. Blue represents uninterrupted “Continuation” of Ryan White Services. In the other scenarios, Ryan White services stop in July 2025. In the “Cessation” scenario (red), viral suppression among Ryan White clients never recovers. In “Prolonged Interruption” (orange), viral suppression recovers from January to December 2029. In “Brief Interruption” (green), viral suppression recovers from January to December 2027.

**Figure S5: State-Level Excess HIV Infections from 2025-2030 If Ryan White Programs are Stopped or Interrupted, *Conservative Secondary Analysis***

|  | **Continuation** | **Cessation** | | **Prolonged Interruption** | | **Brief Interruption** | |
| --- | --- | --- | --- | --- | --- | --- | --- |
| **City** | Number of  Incident Infections | Number of Excess Infections | Relative Excess Infections* | Number of Excess Infections | Relative Excess Infections* | Number of Excess Infections | Relative Excess Infections* |
| Missouri | 2,826 | 1,708 | 61% | 1,326 | 47% | 652 | 23% |
|  | [2,413-3,222] | [1,050-2,490] | [37-86%] | [820-1,923] | [29-67%] | [403-937] | [14-33%] |
| Illinois | 5,105 | 2,513 | 49% | 1,950 | 38% | 981 | 19% |
|  | [4,356-6,203] | [1,604-3,588] | [32-69%] | [1,246-2,759] | [25-53%] | [634-1,373] | [12-27%] |
| Alabama | 3,818 | 1,710 | 45% | 1,313 | 34% | 644 | 17% |
|  | [3,327-4,261] | [1,088-2,444] | [30-62%] | [832-1,873] | [23-47%] | [410-906] | [11-23%] |
| Wisconsin | 1,442 | 609 | 43% | 477 | 33% | 242 | 17% |
|  | [1,145-1,755] | [396-857] | [26-62%] | [310-663] | [21-48%] | [155-339] | [10-25%] |
| Louisiana | 4,545 | 1,640 | 36% | 1,255 | 28% | 605 | 13% |
|  | [3,825-5,288] | [1,075-2,282] | [24-49%] | [824-1,734] | [19-38%] | [395-842] | [9-18%] |
| Florida | 19,222 | 6,243 | 32% | 4,796 | 25% | 2,411 | 13% |
|  | [16,909-22,100] | [3,897-9,138] | [21-46%] | [3,008-6,995] | [16-35%] | [1,519-3,474] | [8-18%] |
| New York | 10,563 | 3,155 | 30% | 2,445 | 23% | 1,233 | 12% |
|  | [9,025-12,200] | [2,012-4,581] | [19-42%] | [1,576-3,512] | [15-33%] | [792-1,737] | [8-16%] |
| Mississippi | 2,565 | 760 | 30% | 587 | 23% | 288 | 11% |
|  | [2,177-3,017] | [491-1,067] | [19-42%] | [378-822] | [15-33%] | [182-402] | [7-16%] |
| Georgia | 12,198 | 3,344 | 27% | 2,601 | 21% | 1,326 | 11% |
|  | [10,687-14,419] | [2,151-4,695] | [18-39%] | [1,672-3,633] | [14-30%] | [850-1,861] | [7-15%] |
| California | 16,519 | 4,432 | 27% | 3,449 | 21% | 1,767 | 11% |
|  | [14,276-19,355] | [2,822-6,371] | [17-39%] | [2,203-4,938] | [13-30%] | [1,113-2,521] | [7-15%] |
| Texas | 24,299 | 4,128 | 17% | 3,179 | 13% | 1,577 | 7% |
|  | [21,391-28,267] | [2,154-6,584] | [9-27%] | [1,666-5,019] | [7-20%] | [829-2,476] | [3-10%] |
| **Medicaid Expansion States** | 39,558 | 13,448 | 34% | 10,425 | 26% | 5,238 | 13% |
|  | [36,801-42,823] | [8,967-18,557] | [22-47%] | [6,950-14,337] | [17-36%] | [3,450-7,184] | [9-18%] |
| **Medicaid Non-Expansion States** | 63,543 | 16,794 | 26% | 12,953 | 20% | 6,487 | 10% |
|  | [59,156-68,774] | [10,852-23,414] | [17-37%] | [8,420-18,121] | [13-29%] | [4,241-9,121] | [7-14%] |
| **Total** | 103,101 | 30,241 | 29% | 23,378 | 23% | 11,725 | 11% |
|  | [97,773-109,048] | [21,095-40,667] | [20-39%] | [16,354-31,275] | [16-30%] | [8,219-15,695] | [8-15%] |
|  |  | 0% \| 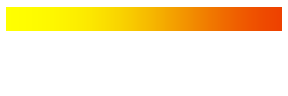 \| 100% | | | | | |

The “Continuation” column gives the mean and 95% credible interval, across 1,000 simulations, projected incident HIV infections from 2025-2030 if Ryan White programs continue uninterrupted. The columns labeled “Number of Excess Infections” give the mean and 95% interval of the absolute number of excess HIV infections expected from 2025-2030 under three scenarios where Ryan White programs are stopped in July 2025. Medicaid non-expansion states are denoted by orange labels, expansion states by purple labels.

**Figure S6: State-Level Relative Excess HIV Infections from 2025-2030 If Ryan White Programs are Stopped or Interrupted, *Conservative Secondary Analysis***

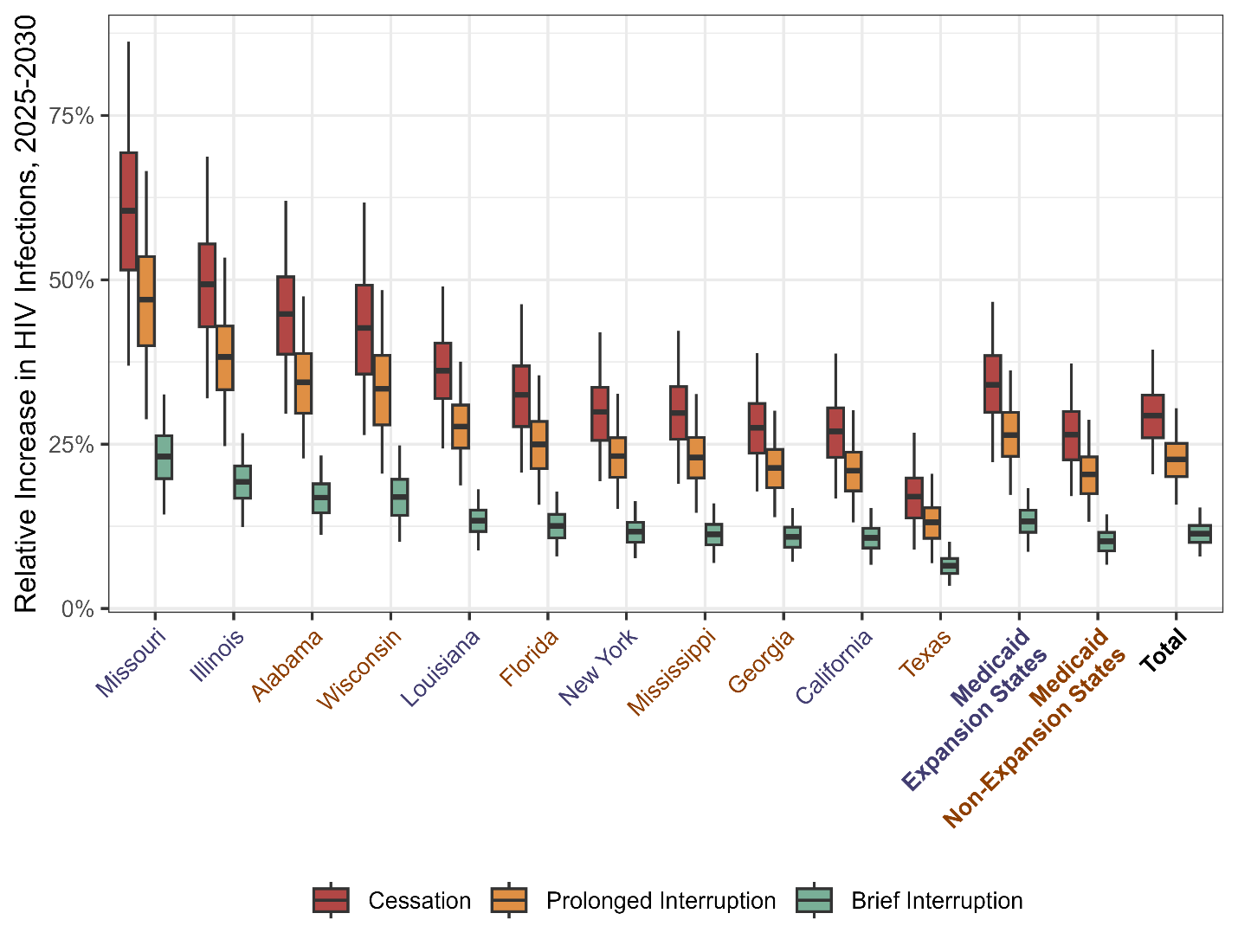

Boxplots display the projected percentage increase in new infections under three scenarios in which all Ryan White services stop in July 2025: “Cessation” (red) – viral suppression among Ryan White clients never recovers; “Prolonged Interruption” (orange) – viral suppression among Ryan White clients recovers from January to December 2029; “Brief Interruption” (green) – viral suppression among Ryan White clients recovers from January to December 2027. The value along the x-axis represents the relative increase in cases vs. a scenario where Ryan White services continue uninterrupted. The dark vertical lines indicate the median projection across 1,000 simulations, the boxes indicate interquartile ranges (IQR), and whiskers cover the 95% credible interval. Medicaid non-expansion states are denoted by orange labels, expansion states by purple labels.

**Figure S7: State-Level Variation in the Projected Relative Increase in HIV Infections if Ryan White Programs End in July 2025**

| 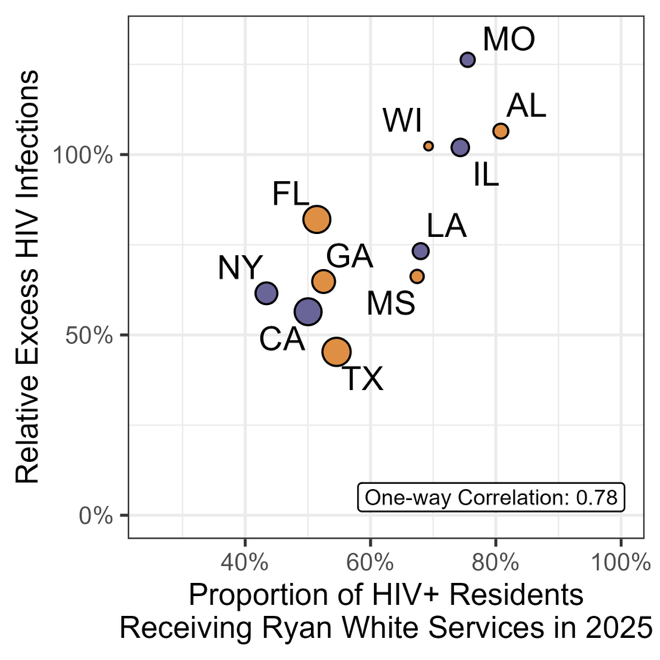 | 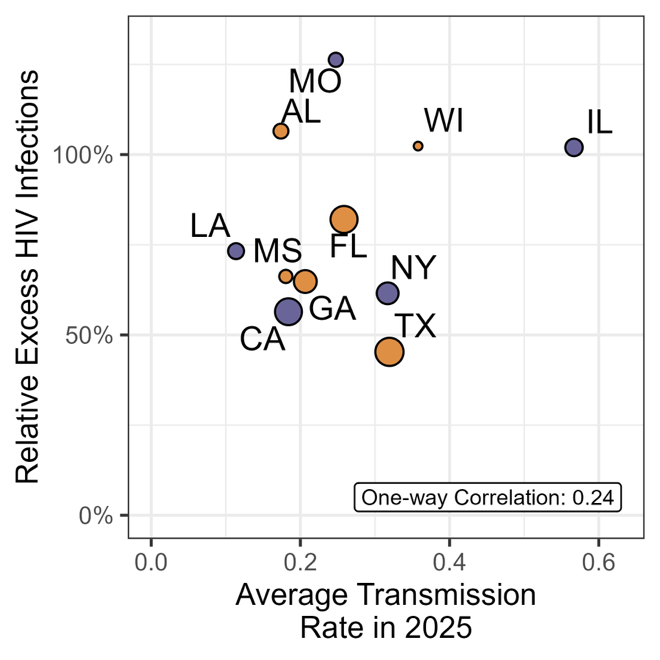 |
| --- | --- |
| 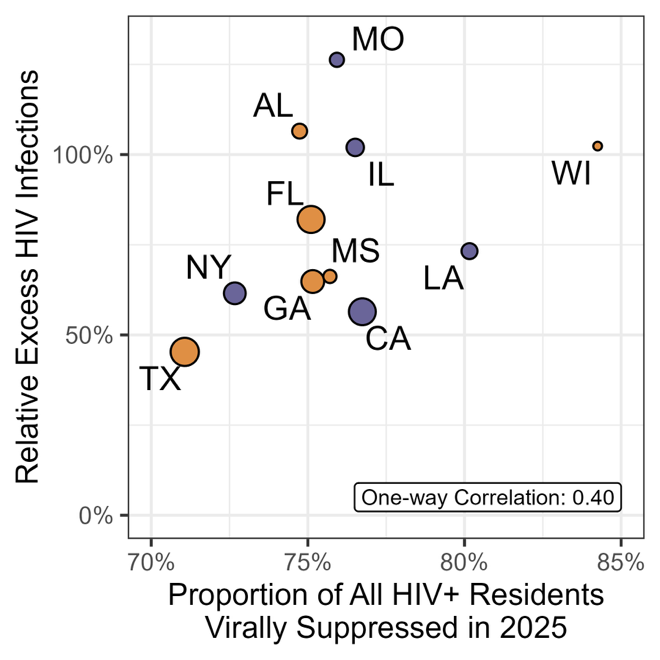 | 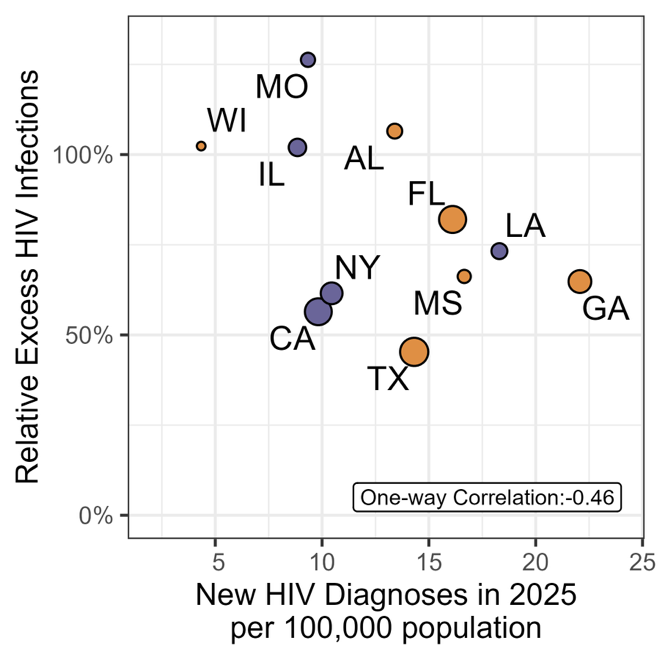 |
| 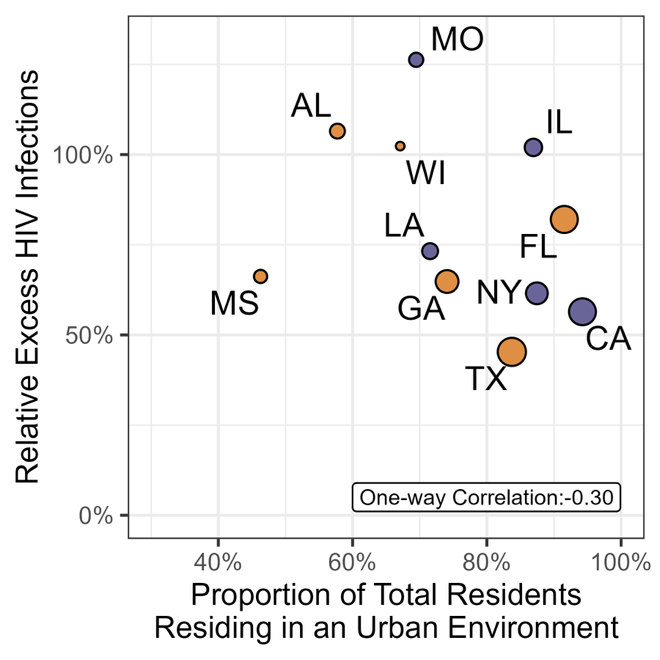 | 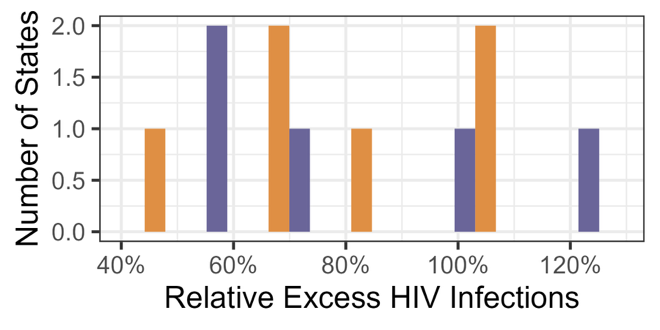  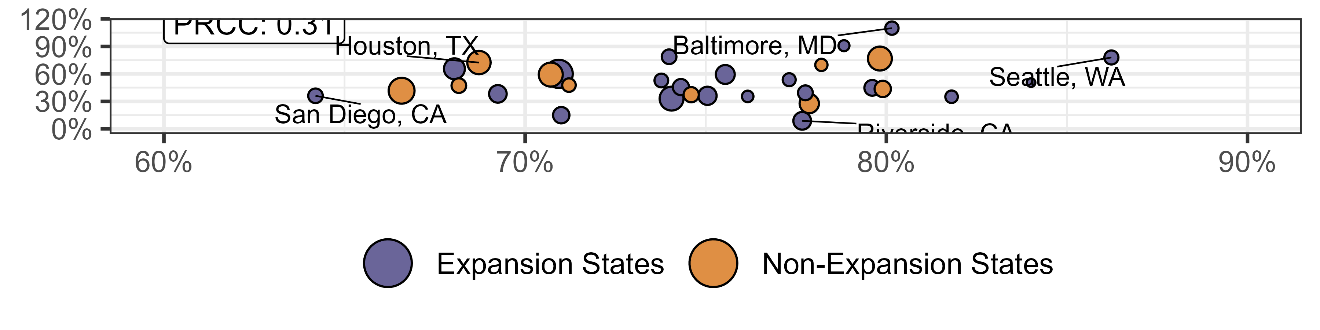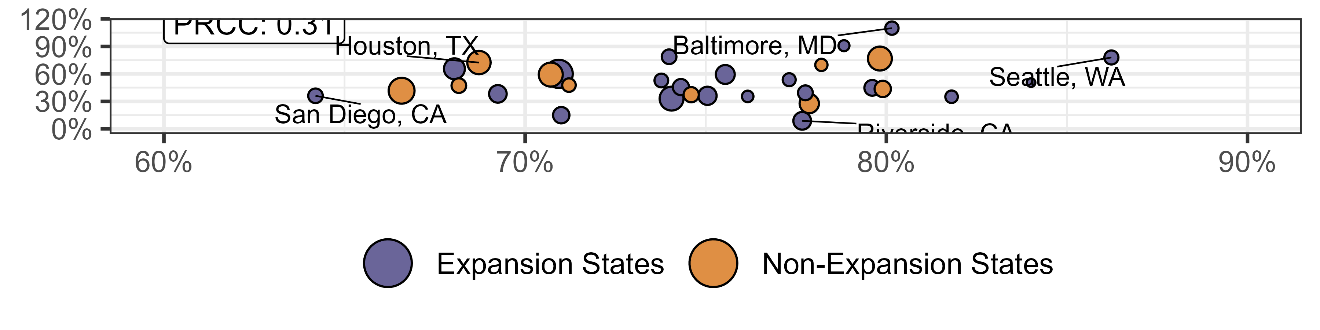 |

Each circle represents one state. The y-axis represents the average relative increase in projected HIV infections from 2025 to 2030 if Ryan White programs end in July 2025 vs. if they continue, averaged across 1,000 simulations. The x-axis represents the average proportion of people with HIV who are Ryan White clients in 2024 (Panel A); average transmission rate (Panel B); average viral suppression among all people with HIV in 2024 (Panel C); average new diagnoses per 100,000 population (Panel D); or average proportion of all residents that belong to an urban environment (Panel E). The size of the circle is proportional to the number of projected new diagnoses in 2024. Panel F shows a histogram of average relative increase in incidence, shaded by Medicaid expansion status. Purple shading indicates Medicaid expansion states; orange indicates non-expansion states. Panels A-E are labeled with the one-way correlation coefficient between each variable and the outcome.

### **Mathematical Details of Model:**

This section describes the mathematical specification of the Ryan White HIV model, which evaluates the impact of program components - the AIDS Drug Assistance Program (ADAP), Outpatient Ambulatory Health Services (OAHS), and other supportive services - on HIV viral suppression. The model differentiates between Medicaid expansion and non-expansion states, and uses survey-derived priors to inform scenario simulations under various scenarios.

#### Population Subgroup Definitions

We model three Ryan White (RW) recipient categories among people with HIV (PWH):

1. ADAP recipients: may or may not receive other RW services.

2. OAHS/non-ADAP recipients: receive OAHS but not ADAP.

3. RW support-only: receive services but neither ADAP nor OAHS services.

#### Mathematical Formulation of Subgroup Proportions

Let
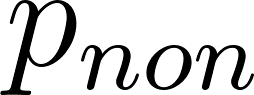
<https://www.codecogs.com/eqnedit.php?latex=p_%7Bnon%7D#0> be the proportion of PWH receiving non-ADAP RW services.

Let
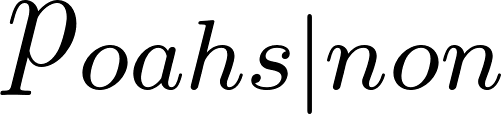
<https://www.codecogs.com/eqnedit.php?latex=p_%7Boahs%7Cnon%7D#0> be the proportion of non-ADAP RW clients receiving OAHS.

Let
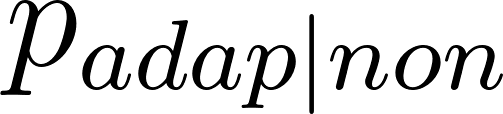
<https://www.codecogs.com/eqnedit.php?latex=p_%7Badap%7Cnon%7D#0> be the proportion of non-ADAP clients who also receive ADAP.

Then:

- Proportion receiving ADAP:
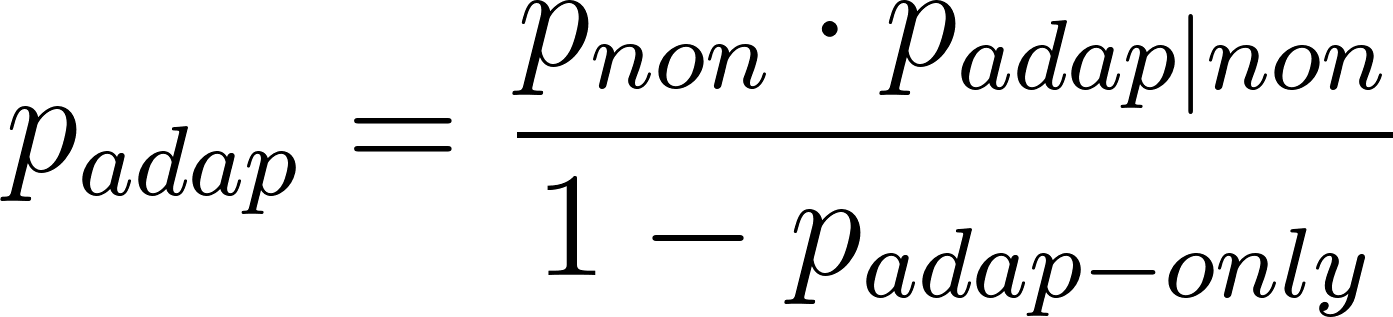
<https://www.codecogs.com/eqnedit.php?latex=p_%7Badap%7D%20%3D%20%5Cfrac%7Bp_%7Bnon%7D%20%5Ccdot%20p_%7Badap%7Cnon%7D%7D%7B1%20-%20p_%7Badap-only%7D%7D#0>

- Proportion receiving OAHS:
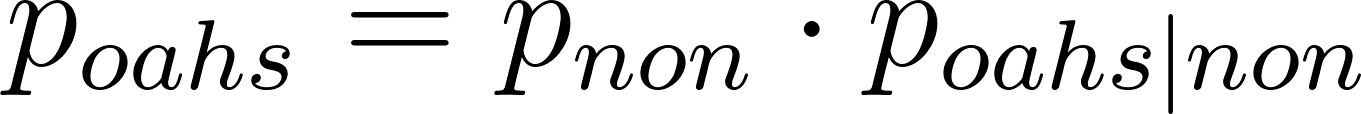
<https://www.codecogs.com/eqnedit.php?latex=p_%7Boahs%7D%20%3D%20p_%7Bnon%7D%20%5Ccdot%20p_%7Boahs%7Cnon%7D#0>

- Proportion receiving supportive services only:
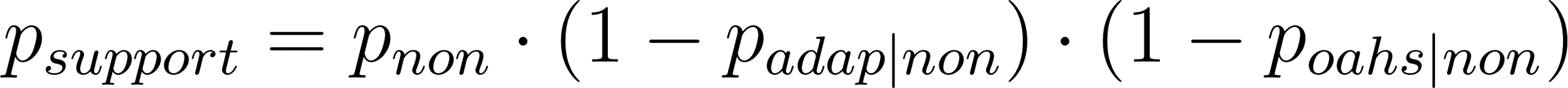
<https://www.codecogs.com/eqnedit.php?latex=p_%7Bsupport%7D%20%3D%20p_%7Bnon%7D%20%5Ccdot%20(1%20-%20p_%7Badap%7Cnon%7D)%20%5Ccdot%20(1%20-%20p_%7Boahs%7Cnon%7D)#0>

- Total RW coverage:
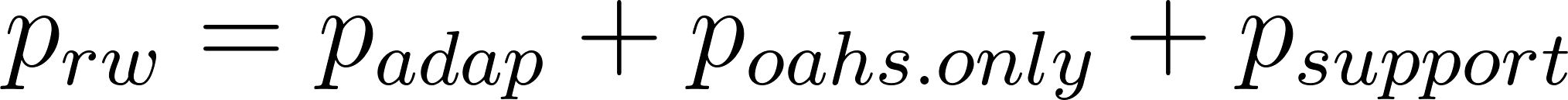
<https://www.codecogs.com/eqnedit.php?latex=p_%7Brw%7D%20%3D%20p_%7Badap%7D%20%2B%20p_%7Boahs.only%7D%20%2B%20p_%7Bsupport%7D#0>

#### Viral Suppression Calculations

Suppression is modeled separately as a proportion of the total number of people with diagnosed HIV. We calculated the suppression among Ryan White client subgroups, conditional on the total viral suppression among all people with HIV in the calibrated base version of the model.

- Suppressed and ADAP Client:
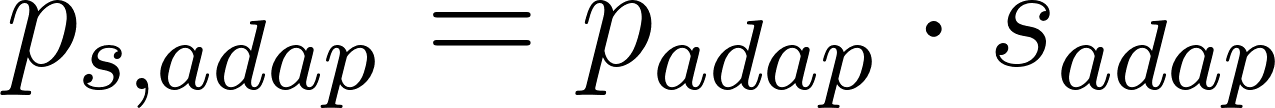
<https://www.codecogs.com/eqnedit.php?latex=p_%7Bs%2Cadap%7D%20%3D%20p_%7Badap%7D%20%5Ccdot%20s_%7Badap%7D#0>

- Suppressed and OAHS Client but not ADAP Client:
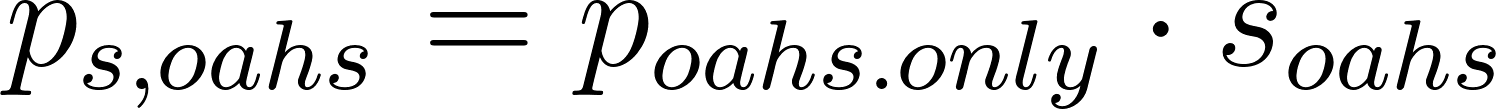
<https://www.codecogs.com/eqnedit.php?latex=p_%7Bs%2Coahs%7D%20%3D%20p_%7Boahs.only%7D%20%5Ccdot%20s_%7Boahs%7D#0>

- Suppressed and receiving supportive services only:
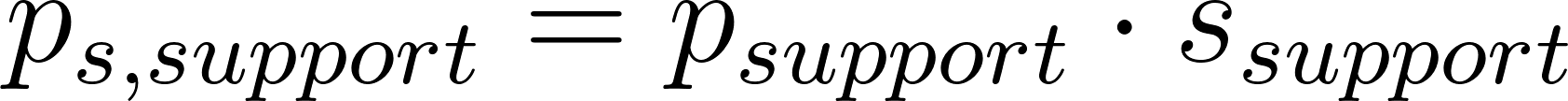
<https://www.codecogs.com/eqnedit.php?latex=p_%7Bs%2Csupport%7D%20%3D%20p_%7Bsupport%7D%20%5Ccdot%20s_%7Bsupport%7D#0>

- Suppressed and not receiving any RW services:
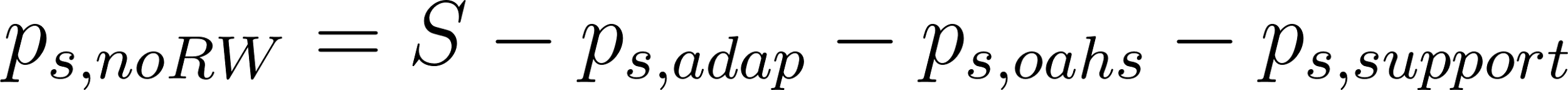
<https://www.codecogs.com/eqnedit.php?latex=p_%7Bs%2CnoRW%7D%20%3D%20S%20-%20p_%7Bs%2Cadap%7D%20-%20p_%7Bs%2Coahs%7D%20-%20p_%7Bs%2Csupport%7D#0>

where
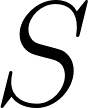
<https://www.codecogs.com/eqnedit.php?latex=S#0> is the total suppression rate among diagnosed PWH.

#### Scenario Effects

We model complete or temporary losses of RW services. For instance, the loss of ADAP in expansion states is:

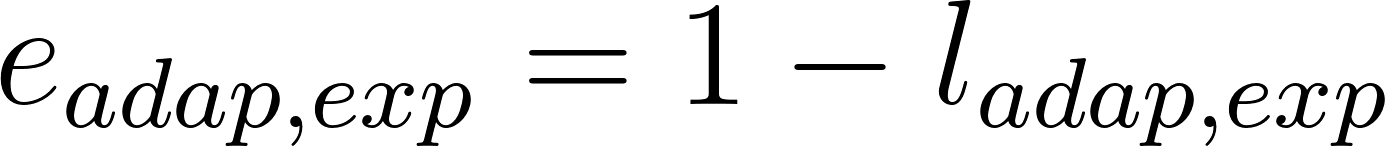
<https://www.codecogs.com/eqnedit.php?latex=e_%7Badap%2Cexp%7D%20%3D%201%20-%20l_%7Badap%2Cexp%7D#0>

where
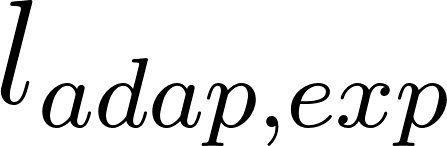
<https://www.codecogs.com/eqnedit.php?latex=l_%7Badap%2Cexp%7D#0> is the loss effect drawn from a kernel density estimate (KDE) of survey data.

Temporary lapses are modeled by changing
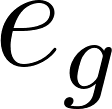
<https://www.codecogs.com/eqnedit.php?latex=e_g#0> over a time interval between interruption and restart.

#### Prior Construction via KDE

Survey responses on expected suppression losses (in %) are transformed using the arcsine-square root:

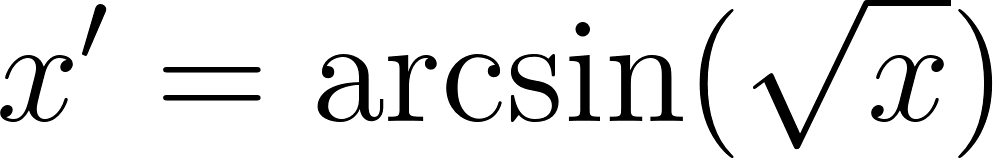
<https://www.codecogs.com/eqnedit.php?latex=x'%20%3D%20%5Carcsin(%5Csqrt%7Bx%7D)#0>

KDE is performed in this transformed space, and values are sampled then inverted as:

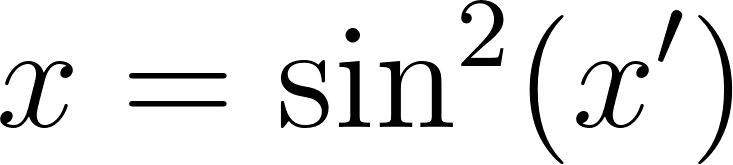
<https://www.codecogs.com/eqnedit.php?latex=x%20%3D%20%5Csin%5E2(x')#0>

This transformation normalizes variance in bounded data and prevents edge effects.
